# Affordable and time-effective high throughput screening of SARS-CoV-2 variants using Denaturing High-Performance Liquid Chromatography analysis

**DOI:** 10.1101/2021.04.27.21256156

**Authors:** Maria Elena Turba, Domenico Mion, Stavros Papadimitriou, Francesca Taddei, Giorgio Dirani, Vittorio Sambri, Fabio Gentilini

## Abstract

**Introduction:** Mutations in the receptor binding domain (RBD) region of SARS-CoV-2 have been shown to impact the infectivity, pathogenicity and transmissibility of new variants of concern (VOC). Even more worrisome, those mutations have the potential of causing immune escape, undermining the population immunity induced by ongoing mass vaccination programs.

**Gap statement:** The massive parallel sequencing techniques have taken a lead role in the detection strategies of the new variants. Nevertheless, they are still cumbersome and labour-demanding. There is an urgent need for novel strategies and techniques aimed at the surveillance of the active emergence and spread of the VOC.

**Aim:** The aim of this study was to provide a quick, cheap and straightforward Denaturing High-Performance Liquid Chromatography (DHPLC) method for the prompt identification of the SARS-CoV-2 VOC.

**Methodology:** Two PCRs were designed to target the RBD region, spanning residues N417 through N501 of the Spike protein. Furthermore, a DHPLC screening analysis was set up. The screening consisted of mixing the unknown sample with a standard sample of a known variant, denaturing at high temperature, renaturing at room temperature followed by a 2-minute run using the WAVE DHPLC system to detect the heteroduplexes which invariably originate whenever the unknown sample has a nucleotide difference with respect to the standard used.

**Results:** The workflow was able to readily detect new variants including the P.1, the B.1.585 and the B.1. 617.2 lineages at a very affordable cost. The DHPLC analysis was robust being able to identify variants even in case of samples with very unbalanced target concentration including those samples at the limit of detection.

**Conclusions:** This approach has the potential of greatly expediting surveillance of the SARS-CoV-2 variants.

## Introduction

The SARS-CoV-2 genome is relatively stable due to the proofreading activity operated by a 3’-to-5’ exoribonuclease (nsp14-ExoN) during replication which reduces the error rate of RNA polymerase 100–1000-fold as compared with other RNA viruses. This confers the capacity of maintaining its 30,000 nt genome to the virus without catastrophic mutational events hampering its integrity [1,2]. Nonetheless, errors still occur in the SARS-CoV-2 genome at a higher rate than eukaryotic cells and, together with high replication rates, allow for the accumulation of mutations in the viral genome including amino acid changes, truncations or the loss of viral proteins [1]. Whenever these changes impact infectivity, pathogenicity and transmissibility, they could lead to higher fitness and undergo positive selection [1,3,4].

The D614G substitution of the Surface protein is the most investigated of the positively selected mutation. This mutation occurred early in the first months of 2020 and became rapidly prevalent worldwide due to the increased infectivity of the strains carrying it. The phenotypic advantage was first evident at the epidemiological level and it was then confirmed at the molecular level using *in-vitro* experiments [5-7].

As the COVID-19 pandemic continues, and vaccination programs expand, however, a novel factor of positive selection has taken place represented by the rapid rise of acquired immunity in the population [8]. As the majority of vaccines exploit the immunogenicity of the Spike protein, due to its pivotal role in binding to the ACE2 cell receptor and entry into the host cell, the strict monitoring of mutations in the domains targeted by the neutralising antibodies should be implemented worldwide [9-11]. In particular, substitutions in the receptor binding domain (RBD) have emerged, and they are of particular concern due to the possibility of being responsible for immune escape. More specifically, the B.1.1.7 (originally identified in the United Kingdom), B.1.351 (Republic of South Africa; RSA), P1 (Brazil), and B.429 (California, USA), B.1.617.1 (India), B.1.617.2 (India) variants carry mutations in the 417, 452, 478 and 484 codons which have strong implications for infectivity and immune evasion [1].

In fact, some of the variants of concern (VOC) which might escape vaccine immunity are being monitored. To this end, an unprecedented sequencing effort has been pushed forward by the implementation of next generation sequencing (NGS) programmes. However, NGS is still cumbersome and very labour intensive. As a consequence, the approach to sequencing any viral genome is still unaffordable and, likely, unwise. Alternatives have been proposed to be integrated with sequencing in order to more effectively monitor the VOC [12,13]. Heteroduplex analysis by Denaturing High-Performance Liquid Chromatography (DHPLC) is a fast, very sensitive and reliable technique to screen nucleic acid variations. It has been used for two decades in many different applications but mainly in detecting cancer somatic mutations due to its extreme analytical sensitivity [14]. When two alleles containing nucleotide variations are denatured by heating and left to renature by cooling as during PCR, in addition to homoduplexes, heteroduplexes due to cross-hybridisation between the mismatched strands are also formed. Therefore, whenever a hotspot for cancer somatic mutations is amplified, any variation may originate heteroduplexes during the PCR amplification. DHPLC discovers DNA variations by separating heteroduplex and homoduplex DNA fragments by ion-pair reverse-phase liquid chromatography [14]. Heteroduplexes and homoduplexes are bound to a stationary phase and eluted by a denaturing gradient at a constant slightly denaturing temperature. Since, heteroduplexes have a lower stability they are eluted before homoduplexes, and they are detected as different peaks modifying the chromatogram shape by ultraviolet absorption.

The aim of this study was to provide a very quick, cheap and straightforward DHPLC method for screening the RBD of SARS-CoV-2 isolates and eventually for readily identifying the SARS-CoV-2 VOC.

## Materials & methods

Positive samples diagnosed with Covid-19 by means of diagnostic reverse transcription polymerase chain reaction (RT-qPCR) (Seegene) underwent DHPLC screening and entire genome sequencing using Oxford Nanopore or Illumina platforms. Furthermore, some samples from External Quality Assessment (EQA) for Molecular Diagnostics were also included in this proof-of-concept study. The overall workflow of the screening proposed is represented in Figure 1.

**Figure 1:**
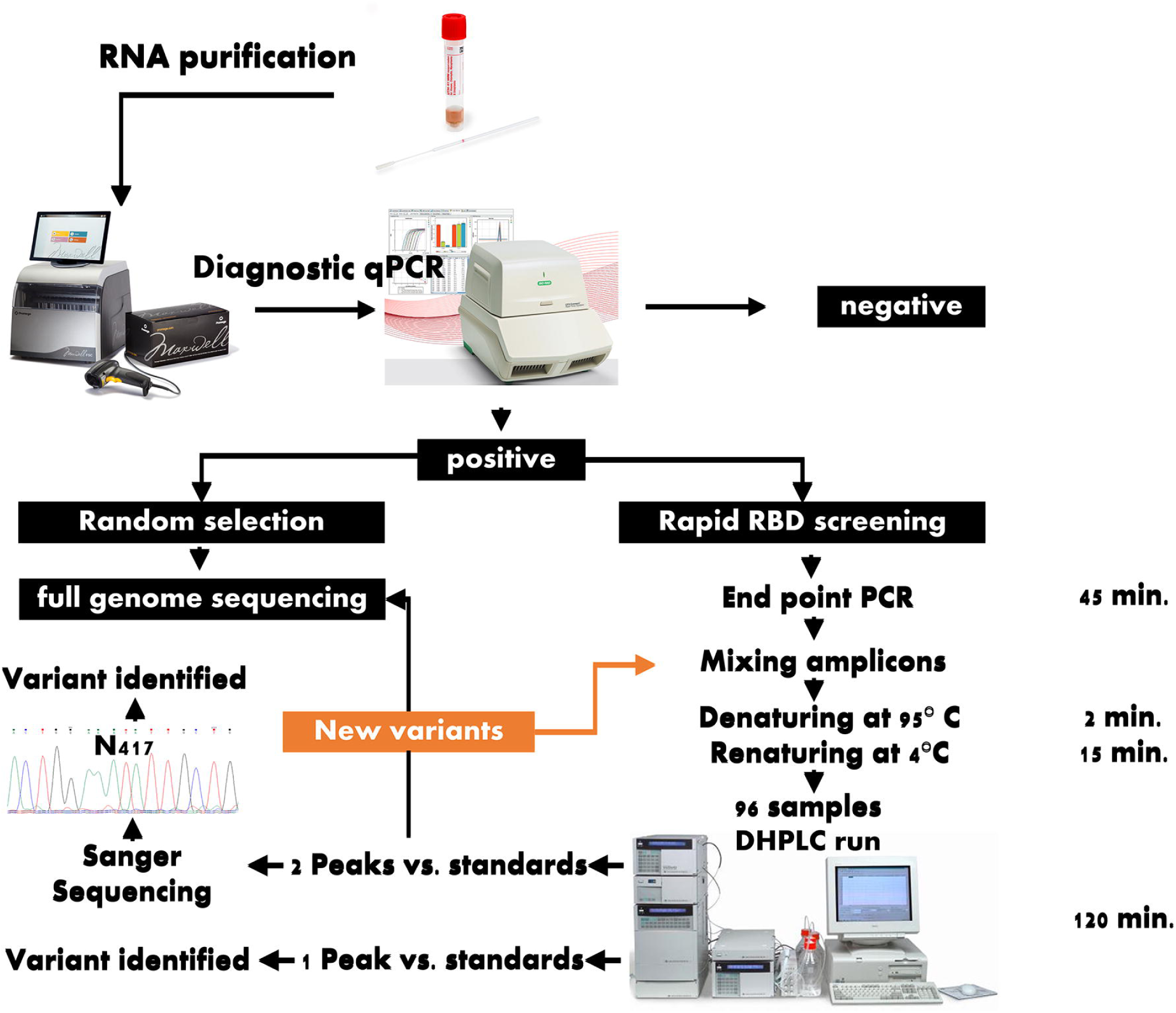
Workflow for the rapid screening of the variants of concern. RBD: SARS-CoV-2 Region Binding Domain; DHPLC: Denaturing High-Performance Liquid Chromatography

For the DHPLC screening, RNA was re-extracted starting from 200 µL of UTM-RT (Copan Italia, Brescia, Italy) samples using a commercial kit (Maxwell® RSC Blood RNA Kit, Promega Italia, Milano, Italy) with an automatic instrument (Maxwell RSC, Promega Italia, Milano, Italy).

The iScript cDNA Synthesis kit (Bio-Rad) was used to reverse transcribe the RNA samples in cDNA (complementary DNA). In brief, 10 µL of RNA were mixed with 4 µL of iScript Reaction mix (containing MMLV RNase H+ reverse transcriptase, dNTPs, oligo(dT)s and random primers), 1 µL of iScript Reverse Transcriptase and nuclease-free water to a final volume of 20 µL and with the following protocol: 5 minutes at 25 °C for priming, 20 minutes at 46 °C for reverse transcription and 1 minute at 95 °C for reverse transcriptase inactivation.

cDNA thereby obtained was consequently used as a template for PCR.

The PCR reactions for amplifying two sub-regions within the RBD of the S gene were designed using the Primer Quest web design tool (https://eu.idtdna.com/Primerquest/Home/Index). The PCR reactions were designed keeping the amplicon approximately 200-300 bp long and by including the 417 and 452 codons in the RBD_1 PCR, the 452, 478, 484 and 501 codons in the RBD_2 PCR. The PCR primers were F_Var417_NCov19, 5’ TAAGTGTTATGGAGTGTCTCCTACTA 3’, R_Var417_NCov19, 5’ GGTTTGAGATTAGACTTCCTAAACAATC 3’, F_Var484_501_NCov19, 5’ TCTTGATTCTAAGGTTGGTGGTAA 3’ and R_Var484_501_NCov19, 5’ AGTTGCTGGTGCATGTAGAA 3’.

Briefly, the PCR reactions consisted of 1X Buffer (Phusion HF Buffer, Life Technologies), 0.2 mM MgCl_2_, 350 nM each of the forward and reverse primers, deoxynucleotide triphosphates (dNTPs) 250 µM, 0.4 U of Phusion Taq DNA polymerase (ThermoFisher, Milan, Italy), 2 µL of cDNA template brought up to a final volume of 20 µL with molecular biology grade water. The cycling programme for both the conventional PCRs consisted of the following steps: 98°C for 30 s, 40 cycles at 98°C for 5 s, 63°C for 5 s and 72°C for 10 s, and a final elongation step at 72° C for 10 min.

### DHPLC

In this study, heteroduplexes if any, were generated by mixing the amplicons of the above-described end-point PCRs of a standard (reference) sample and a test (unknown) sample and by denaturing the mixed fragments at 95°C for 2 min, and then allowing to renature at room temperature for 15 min. Each test sample was then run as such and mixed with the reference (standard) on an automated DHPLC apparatus (WAVE® DHPLC system, ADS Biotec, Omaha NE, USA) equipped with a proprietary column (DNASep®, ADS Biotec, Omaha NE, USA) which uses alkylated non-porous polystyrene-divinylbenzene copolymer hydrophobic beads for high performance nucleic acid separations. Separation of products was carried out by a mobile phase obtained by continuously mixing buffer B (0.1 mol/L triethylammonium acetate–TEAA with 25% acetonitrile) to buffer A (0.1 mol/L triethylammonium acetate–TEAA), according to a gradient (Table Y) calculated by the Navigator™ Software (ADS Biotec, Omaha NE, USA) and experimentally confirmed.

Likewise, the optimal oven temperatures for heteroduplex separation were determined using Navigator™ Software which gave a computer□assisted determination of the melting profile and analytical conditions for each fragment which were then experimentally verified. In this study, a partially denaturing temperature of 55.5°C was used for the DHPLC screening of the N417T/N mutation in RBD_1 amplicon and a partially denaturing temperature of 56.6° C for the E484K and N501Y mutations in the RBD_2 amplicon.

In the DHPLC system, amplicons are screened for chromatogram shape and in particular for the presence or not of more than one peak with respect with the reference control. Since, the method is not quantitative but qualitative, it is not relevant the peak height and thereafter the peak quantitation. Conversely, the inherent impressive analytical sensitivity demonstrated by hundreds of papers to be as low as 1%, allow for detecting an heteroduplex peak even if the amount of mixed amplicons are not quantified and normalized.

Of course, in order to readily identify additional peaks unveiling a nucleotide variation with respect to the standard used, the test sample should be adequately amplified. To avoid the needs of quantifying and normalizing the nucleic acid of the variants and hence to speed up the workflow, a serial dilution experiment was carried out by spiking in human saliva viral mRNA of a B1.617.2 variant sample serially diluted 1:10 in molecular biology grade water from 3.5.0×10^8^ to 3.5×10^3^ copies/mL. MRNA was then purified from the saliva samples spiked with the viral mRNA and the mRNA retrotranscribed according with the above-mentioned methods, The cDNA samples were then PCR amplified, checked for the presence of an amplification band and then tested by DHPLC to assess the last dilution yielding a distinct heteroduplex peak.

Before each run, the column was prepared according with the manufacturer’s instruction. In particular, the Wave Low-range mutation control standard and Wave High-range mutation control standard were used to check the apparatus before each run. One reference control was included in each assay run as well as a blank sample. To optimise the workflow, as reference control should be used the more prevalent variant identified in the lab in a given period. In this study, either the B.1 or B.1.177 or B.1.617.2 were used as standard.

The data analysis was carried out using Navigator™ Software. Following the DHPLC screening, the same amplicons were purified and sequenced using an ABI310 automated sequencer (ThermoFisher, Milan, Italy). All the variants used in the study had already been fully sequenced using a modified Artic protocol (unpublished) by using Nanopore Sequencing (Oxford Nanopore Technologies, Oxford, UK) or Illumina platform (Illumina Italy, Milan, Italy).

According with the workflow in Fig. 1, all positive samples should undergo to the screening.

## Results

The end-point PCR assays showed discrete bands without smearing and additional non-specific bands (Figure 2).

**Figure 2:**
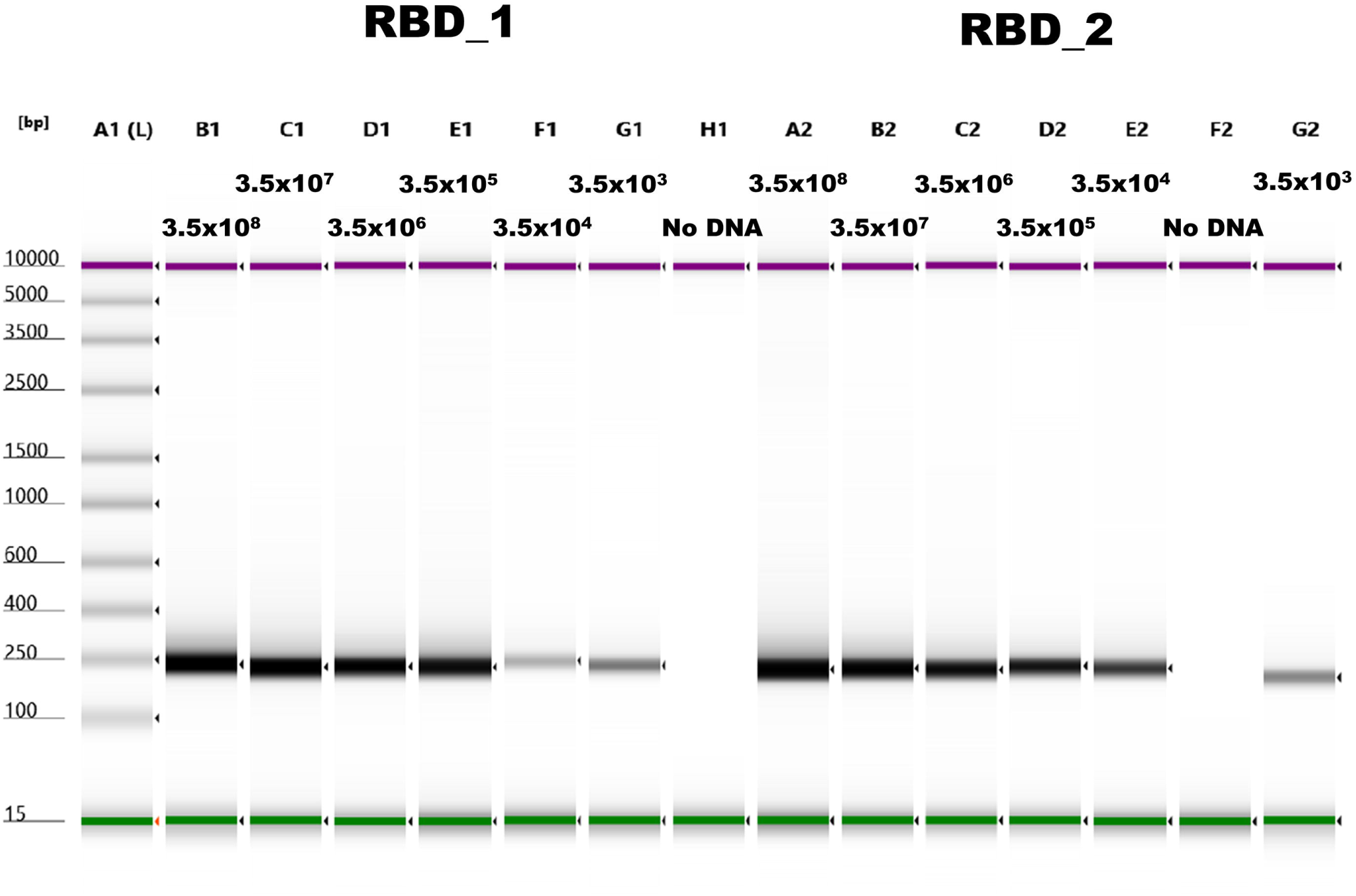
Gel image of the electrophoresis analysis of the RBD_1 and RBD_2 end-point PCR. A1 lane shows the sizing ladder. Purple and green bands represent the upper and lower markers, respectively. The expected viral load expressed as copies/mL are indicated above each lane. Discrete sharp bands without smearing and additional non-specific bands are clearly identifiable in all cases. RBD: Region Binding Domain of SARS-CoV-2.

Overall, the DHPLC identified the B.1.1.7 variant using the B.1 as a reference (Figure 3) and then using the B.1.1.7 as a reference, the DHPLC assay identified the emerging VOC P.1, B.1.585 (Figure 4) and the most recently emerging B.1.617.2 (Figure 5).

**Figure 3:**
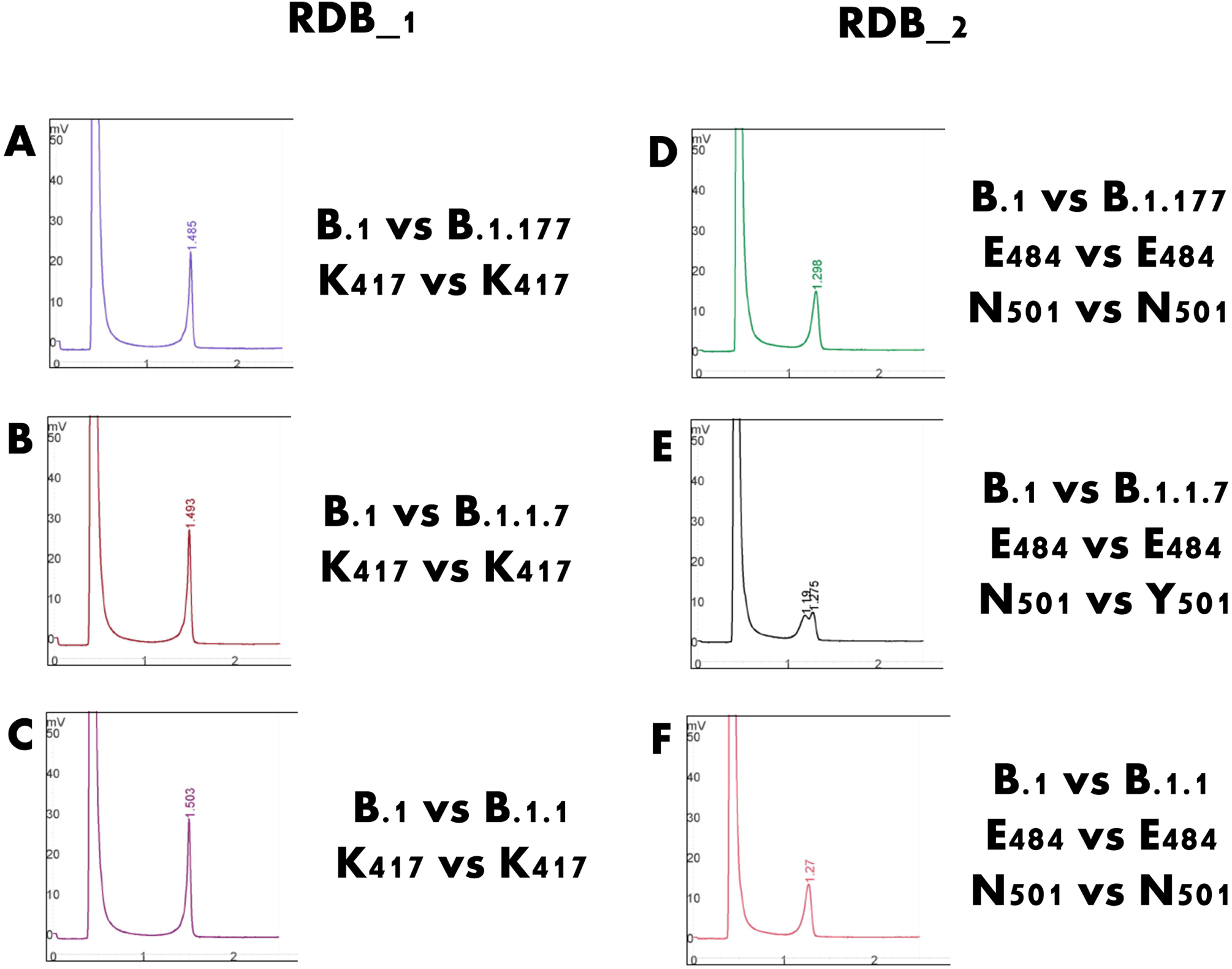
Detection of Y501 residue (B.1.1.7 variant) throughout heteroduplex detection between the region binding domain Target 2 (RBD_2) amplified from a B.1 strain and a B.1.1.7 strain. The mixing, denaturing at 95°C and renaturing at room temperature between two mutations causes a heteroduplex formation detected by the Wave Denaturing High-Performance Liquid Chromatography system, such as peak splitting in E. A, B, C, D and F represent the typical appearance of homoduplexes.

**Figure 4:**
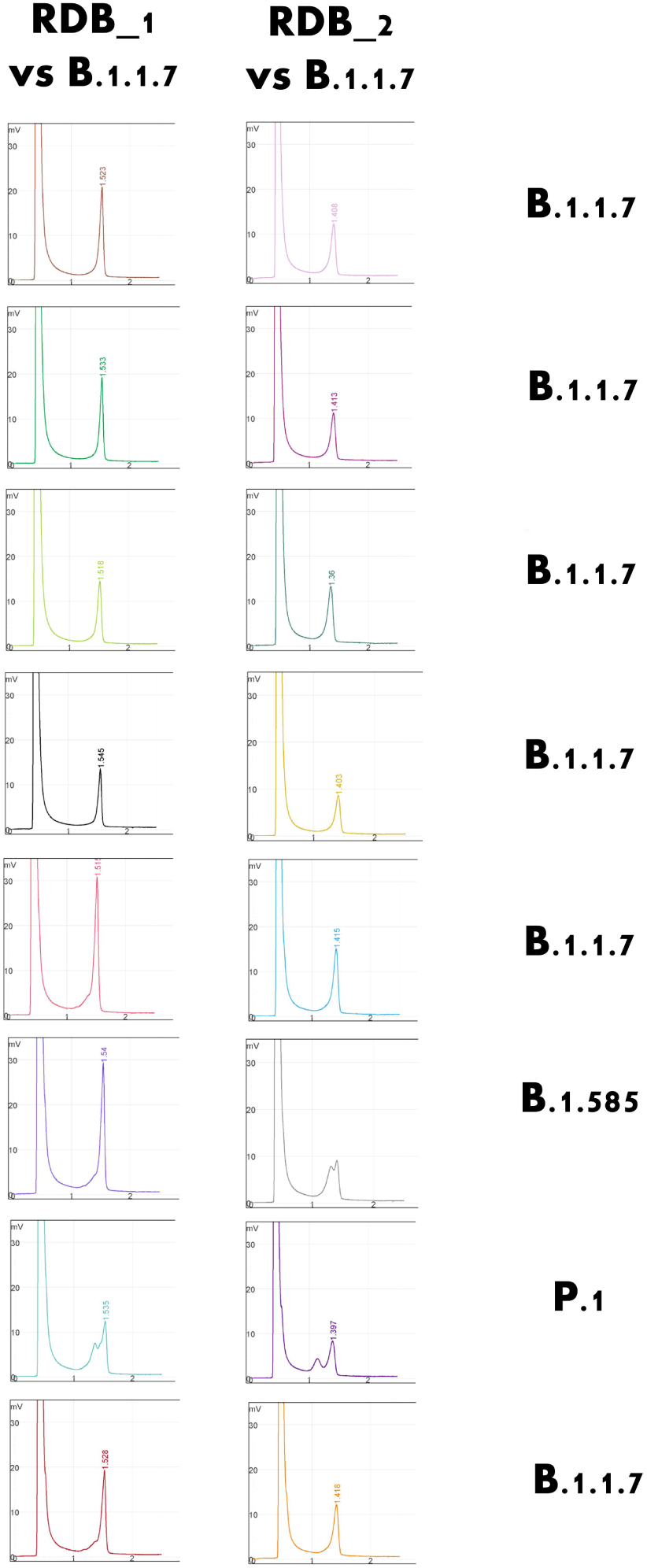
Selected example of the detection of rare variants of concern using the Denaturing High-Performance Liquid Chromatography system. Using the B.1.1.7 strain as a standard, the rarer B.1.585 (Nigerian) and P.1. (Brazilian) variants could be detected and confirmed.

**Figure 5:**
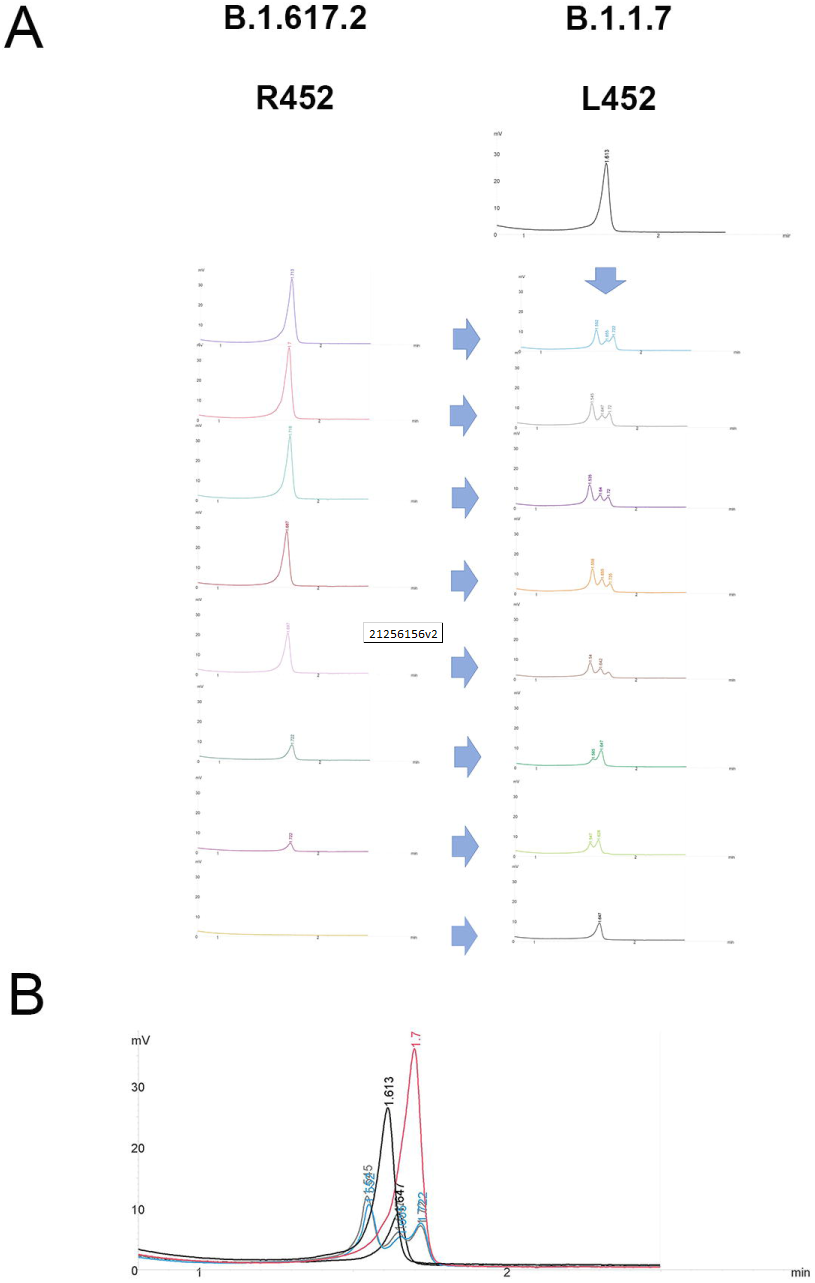
Denaturing High-Performance Liquid Chromatography. A) dilutional experiment to verify the effect of unbalanced concentration of amplicons. Example of the RBD_1 target screened by mixing amplicons obtained from samples containing different viral loads (B.1.617.2) with the sample reference sample (B.1.1.7). The two variants present a L452R amino acid variant. The samples containing heteroduplexes can be readily identified at any initial viral load. B) Higher magnification of overlapping chromatograms of reference sample (B.1.1.7; red curve) and test (B.1.617.2; black curve) samples as such and mixed each other to originate heteroduplexes appearing as multiple peaks modifying the chromatogram shape (cyan and grey curves). Also, the no-DNA control mixed with the reference samples is shown as lower light grey curve containing a single peak)

Even samples with very low viral load, very close to the limit of detection of the diagnostic PCR, could be amplified and, remarkably, they could be readily screened by DHPLC (Figure 5). The method allowed to identify heteroduplexes carried out by mixing the B.1.117 and the emerging B.1.617.2 variant even at the 7 copies/reaction concentration.

## Discussion

DHPLC exploits the different retention times of homoduplexes and heteroduplexes in order to detect their multiple retention peaks. A sample of a known sequence was used as a standard and was mixed with unknown samples. Whenever more than one retention peak was evident at DHPLC, it indicated that a mismatch, and hence a mutation, existed between the standard and the sample. To achieve adequate separation of the elution peaks, DHPLC are suitable to screen PCR fragments with a relatively limited length ranging between 150 and 400 bp [14]. To almost cover the entire RBD of the Spike protein, two PCRs were designed, and the respective amplicons were screened for mutations using DHPLC. As an inherent limitation of the technique, the need of further including viral targets would need additional PCR reaction. On the other hand, the setup of the PCR assays was straightforward. To obtain sharp and easily interpretable peaks at DHPLC analysis, PCR reactions should avoid the formation of nonspecific amplicons and smears (Figure 2). The high-fidelity Phusion Taq polymerase is well-suited for this purpose; furthermore, the PCR reactions using Phusion Taq are very rapid in reducing the PCR step. It should also be pointed out that the Phusion buffer does not contain any detergent which would be detrimental for downstream DHPLC analysis.

In this study involving DHPLC analysis, the B.1 strain isolated in March 2020 carrying the K417, E484 and N501 residues in the RBD_1 and RBD_2 amplicons were first used. Using this strain, the prevalent Y501 [15] could be identified (Figure 2) and used as a standard for subsequent analysis which allowed detecting the P.1 and B1.585 variants (Figure 3), as confirmed by Sanger Sequencing of the amplicons and whole genome sequencing using Oxford Nanopore (Oxford Nanopore Technologies). Finally, the method allowed to identify the B.1.617.2.

Overall, the process was able to be completed in a few hours. Potentially, all new positives would be able to be screened on the same day of diagnosis using a dedicated thermal cycler and the WAVE® DHPLC system (Figure 1). Besides the cost of the additional PCR reaction, an additional reagent cost for each DHPLC analysis would be 1.40 Euros/amplicon. This amounts to 2.80 Euros/sample when considering both RBD_1 and RBD_2 targets.

The Spike protein has different hotspots of mutation and deletion; the most likely candidates for immune escape are those within the RBD, such as K417N/T, E484K, and N501Y; N439K, L452R, Y453F, S477N and T478K, are also likely to have a negative impact on the effectiveness of the vaccination [1, 5, 16-18]. Alarmingly, a convergent evolution has led to diverse groupings of those mutations among different clades [10]. Using known sequenced strains, DHPLC is able to rapidly screen a large number of samples, establishing whether they share the same combination of mutations in the RBD or eventually not. In the latter case, additional combinations may be rapidly ascertained in a few minutes by mixing and denaturing at high temperature, allowing renaturing and running in DHPLC with amplicons from known mutation combination. Other possible targets could also be added to the analysis whenever their role in vaccination escape in other domains emerges [19,20]. The flexible, cost-effective streamlined approach herein described, which integrates DHPLC with Sanger sequencing and eventually NGS, should be considered to implement surveillance of the VOC.

Compared to other methods like multiplex real-time PCR genotyping [21], multiplex ARMS PCR [22] the assay does not require complicated procedures for establishment or maintaining the analytical performance, or cumbersome technical replicates. Moreover, these latter techniques are strictly sequence specific and can only find known variants with the aim of establishing their prevalence. On the contrary, the DHPLC screening may detect all the variants, either the known or the still unknown ones.

The very rapid checking of the presence of the PCR amplicon of the test sample using a fragment analyser or an agarose gel guarantee that the presence of heteroduplex and, hence, the presence of variation with respect to the standard used readily prompt the presence of a different variation in the RBD region of Sars-CoV2.

## Data Availability

All data are available at request from the Authors

## Authors’ contributions

Conceptualization, FG, MET; formal analysis, DM, FT, GD; data curation, FG, MET, DM, FT, GD; writing—original draft preparation, FG, MET; writing—review and editing, SP and VS; validation, DM, FT, GD, and VS; supervision, FG and SP; project administration, MET; funding acquisition, VS. All authors have read and agreed to the published version of the manuscript

## Ethical approval

The study was conducted according to the guidelines of the Declaration of Helsinki, and approved by the Institutional Review Board of AUSL Romagna under the protocol code “COVdPCR of 07/02/2020 (it includes appropriate approvals or waivers).

## Consent for publication

Not applicable. Informed Consent Statement: The samples included in this study were sent to the Unit of Micro-biology, Greater Romagna Area Hub Laboratory, Cesena, Italy, for routine diagnostic purposes and the laboratory data results were reported as answer to a clinical suspicion. As such, informed consent from patients was not required. Each sample included was preventively anonymized.

## Acknowledgements

Some viral strain included in the study had been investigated within the interlaboratory “INSTAND e.V. “external proficiency testing program.

## Reference

1. Plante JA, Liu Y, Liu J, Xia H, Johnson BA, et al. Spike mutation D614G alters SARS-CoV-2 fitness. Nature 2021;592:116–121. doi: 10.1038/s41586-020-2895-3.

2. Triggle CR, Bansal D, Ding H, Islam MM, Farag EABA, et al. A Comprehensive Review of Viral Characteristics, Transmission, Pathophysiology, Immune Response, and Management of SARS-CoV-2 and COVID-19 as a Basis for Controlling the Pandemic. Front Immunol 2021;12:631139. doi: 10.3389/fimmu.2021.631139.

3. Giovanetti M, Benedetti F, Campisi G, Ciccozzi A, Fabris S, et al. Evolution patterns of SARS-CoV-2: Snapshot on its genome variants. Biochem Biophys Res Commun 2021;538:88–91. doi: 10.1016/j.bbrc.2020.10.102.

4. Li Q, Wu J, Nie J, Zhang L, Hao H, et al. The Impact of Mutations in SARS-CoV-2 Spike on Viral Infectivity and Antigenicity. Cell 2020;182:1284-1294.e9. doi: 10.1016/j.cell.2020.07.012.

5. Groves DC, Rowland-Jones SL, Angyal A. The D614G mutations in the SARS-CoV-2 spike protein: Implications for viral infectivity, disease severity and vaccine design. Biochem Biophys Res Commun 2021;538:104–107. doi: 10.1016/j.bbrc.2020.10.109.

6. Jackson CB, Zhang L, Farzan M, Choe H. Functional importance of the D614G mutation in the SARS-CoV-2 spike protein. Biochem Biophys Res Commun 2021;538:108–115. doi: 10.1016/j.bbrc.2020.11.026.

7. Plante JA, Mitchell BM, Plante KS, Debbink K, Weaver SC, et al. The variant gambit: COVID-19’s next move. Cell Host Microbe 2021:S1931-3128(21)00099-8. doi: 10.1016/j.chom.2021.02.020.

8. Jung J. Preparing for the Coronavirus Disease (COVID-19) Vaccination: Evidence, Plans, and Implications. J Korean Med Sci. 2021 Feb 22;36(7):e59. doi: 10.3346/jkms.2021.36.e59.

9. Cobey S, Larremore DB, Grad YH, Lipsitch M. Concerns about SARS-CoV-2 evolution should not hold back efforts to expand vaccination. Nat Rev Immunol 2021:1–6. doi: 10.1038/s41577-021-00544-9.

10. Focosi D, Maggi F. Neutralising antibody escape of SARS-CoV-2 spike protein: Risk assessment for antibody-based Covid-19 therapeutics and vaccines. Rev Med Virol 2021 Mar 16. doi: 10.1002/rmv.2231.

11. Gómez CE, Perdiguero B, Esteban M. Emerging SARS-CoV-2 Variants and Impact in Global Vaccination Programs against SARS-CoV-2/COVID-19. Vaccines (Basel) 2021;9:243. doi: 10.3390/vaccines9030243.

12. Bal A, Destras G, Gaymard A, Stefic K, Marlet J, et al. Two-step strategy for the identification of SARS-CoV-2 variant of concern 202012/01 and other variants with spike deletion H69-V70, France, August to December 2020. Euro Surveill 2021;26(3):2100008. doi: 10.2807/1560-7917.ES.2021.26.3.2100008.

13. Banada P, Green R, Banik S, Chopoorian A, Streck D, et al. Simple RT-PCR Melting temperature Assay to Rapidly Screen for Widely Circulating SARS-CoV-2 Variants. medRxiv. 2021 Mar 8:2021.03.05.21252709. doi:10.1101/2021.03.05.21252709. Preprint.

14. Xiao W, Oefner PJ. Denaturing high-performance liquid chromatography: A review. Hum Mutat 2001;17:439–474. doi: 10.1002/humu.1130.

15. Kirby T. New variant of SARS-CoV-2 in UK causes surge of COVID-19. Lancet Respir Med 2021;9:e20–e21. doi: 10.1016/S2213-2600(21)00005-9.

16. Garcia-Beltran WF, Lam EC, St Denis K, Nitido AD, Garcia ZH, et al. Multiple SARS-CoV-2 variants escape neutralization by vaccine-induced humoral immunity. Cell. 2021 Mar 12:S0092-8674(21)00298-1. doi: 10.1016/j.cell.2021.03.013.

17. Karim SSA. Vaccines and SARS-CoV-2 variants: the urgent need for a correlate of protection. Lancet 2021;397:1263–1264. doi: 10.1016/S0140-6736(21)00468-2.

18. Knoll MD, Wonodi C. Oxford-AstraZeneca COVID-19 vaccine efficacy. Lancet 2021;397:72–74. doi: 10.1016/S0140-6736(20)32623-4.

19. McCarthy KR, Rennick LJ, Nambulli S, Robinson-McCarthy LR, Bain WG, et al. Recurrent deletions in the SARS-CoV-2 spike glycoprotein drive antibody escape. bioRxiv. 2020:2020.11.19.389916. (Preprint)

20. Pereira F. SARS-CoV-2 variants combining spike mutations and the absence of ORF8 may be more transmissible and require close monitoring. Biochem Biophys Res Commun 2021;550:8–14. doi: 10.1016/j.bbrc.2021.02.080.

21. Vogels CBF, Breban MI, Ott IM, Alpert T, Petrone ME, et al. Multiplex qPCR discriminates variants of concern to enhance global surveillance of SARS-CoV-2. PLoS Biol 2021;19(5):e3001236. doi: 10.1371/journal.pbio.3001236.

22. Islam MT, Alam ARU, Sakib N, Hasan MS, Chakrovarty T, et al. A rapid and cost-effective multiplex ARMS-PCR method for the simultaneous genotyping of the circulating SARS-CoV-2 phylogenetic clades. J Med Virol 2021;93:2962–2970. doi: 10.1002/jmv.26818.

